# Effects of SARS-CoV-2 P.1 introduction and the impact of COVID-19 vaccination on the epidemiological landscape of São José Do Rio Preto, Brazil

**DOI:** 10.1101/2021.07.28.21261228

**Authors:** Cecília Artico Banho, Lívia Sacchetto, Guilherme Rodrigues Fernandes Campos, Cíntia Bittar, Fábio Sossai Possebon, Leila Sabrina Ullmann, Beatriz de Carvalho Marques, Gislaine Ceslestino Dutra da Silva, Marília Mazzi Moraes, Maisa Carla Pereira Parra, Andreia Francesli Negri, Ana Carolina Boldrin, Michela Dias Barcelos, Thayza M. I. L. dos Santos, Bruno H. G. A. Milhim, Leonardo Cecílio da Rocha, Fernanda Simões Dourado, Andresa Lopes dos Santos, Victoria Bernardi Ciconi, Caio Patuto, Alice Freitas Versiani, Rafael Alves da Silva, Edoardo Estevam de Oliveira Lobl, Victor Miranda Hernandes, Nathalia Zini, Carolina Colombelli Pacca, Cássia Fernanda Estofolete, Helena Lage Ferreira, Paula Rahal, João Pessoa Araújo, Jamie A. Cohen, Cliff C. Kerr, Benjamin M. Althouse, Nikos Vasilakis, Mauricio Lacerda Nogueira

**Affiliations:** Department of Dermatological, Infectious, and Parasitic Diseases, Faculdade de Medicina de São José do Rio Preto, São José do Rio Preto, São Paulo, Brazil; Department of Biology, São Paulo State University, Institute of Biosciences, Languages and Exact Sciences, São José do Rio Preto, São Paulo, Brazil; Biotechnology Institute, São Paulo State University, Botucatu, São Paulo, Brazil; Epidemiological Surveillance Department, São José do Rio Preto, São Paulo, Brazil; Faceres Medical School, São José do Rio Preto, São Paulo, Brazil; Department of Veterinary Medicine, FZEA-USP, University of São Paulo, Pirassununga, São Paulo, São Paulo, Brazil; Institute for Disease Modeling, Global Health Division, Bill & Melinda Gates Foundation, Seattle, WA, USA; University of Washington, Seattle, Washington, United States of America; New Mexico State University, Las Cruces, New Mexico, United States of America; Department of Pathology, University of Texas Medical Branch, Galveston, Texas, United States of America; Sealy Center for Vector-Borne and Zoonotic Diseases, University of Texas Medical Branch, Galveston, Texas, United States of America; Center for Biodefense and Emerging Infectious Diseases, University of Texas Medical Branch, Galveston, United States of America; Center for Tropical Diseases, University of Texas Medical Branch, Galveston, Texas, United States of America; Institute for Human Infection and Immunity, University of Texas Medical Branch, Galveston, Texas, United States of America

**Keywords:** P.1 lineage, genomic surveillance, variant of concern, clade replacement, vaccine effect

## Abstract

The emergence of the new Brazilian variant of concern, P.1 lineage (Gamma), raised concern about its impact on the epidemiological profile of COVID-19 cases due to its higher transmissibility rate and immune evasion ability. Using 272 whole-genome sequences combined with epidemiological data, we showed that P.1 introduction in São José do Rio Preto, São Paulo, Brazil, was followed by the displacement of eight circulating SARS-CoV-2 variants and a rapid increase in prevalence two months after its first detection. Our findings support that the P.1 variant is associated with an increase in mortality risk and severity of COVID-19 cases in younger aged groups, which corresponds to the unvaccinated population at the time. Moreover, our data highlight the beneficial effects of vaccination indicated by a pronounced reduction of severe cases and deaths in immunized individuals, reinforcing the need for rapid and massive vaccination.

## Introduction

The severe acute respiratory syndrome coronavirus 2 (SARS-CoV-2) has affected millions of lives worldwide ^1^, and with its rapid spread and evolution, variants have emerged and continue to emerge globally ^2^ . Currently, SARS-CoV-2 variants that present a threat to global health are classified as Variants of Interest (VOIs) and Variants of Concern (VOCs) ^3^ . To date, all VOCs are descendants of the SARS-CoV-2 strain containing a S: D614G mutation, which is associated with increased viral fitness through enhancement of viral load in the upper respiratory tract, favoring viral spread and transmission ^4,5^ . As of July 2021, four reported lineages had been defined as VOCs: B.1.1.7 (Alpha), first detected in the United Kingdom (U.K.) in December 2020, has been associated to enhanced transmissibility, higher severity of disease ^6,7^, and lower neutralization potential by vaccine and convalescent sera ^8^ ; B.1.351 (Beta), a variant first detected in South Africa, has been associated with an increased risk of transmission and reduced neutralization by monoclonal antibody therapy, convalescent sera, and post-vaccination sera ^9–12^ ; P.1 (Gamma), a Brazilian variant derived from the B.1.1.28 lineage, shares some critical mutations in the Spike protein with B.1.351 ^13^ ; and B.1.617 (Delta), detected in India at the end of 2020, has spread and replaced the B.1.1.7 in the U.K. and showed significantly reduced neutralizing antibody activity compared with the wild-type strain and other VOCs ^14^ .

The Brazilian VOC, P.1 lineage, emerged in Manaus city, the capital of Amazonas state, in November 2020 ^15^ and rapidly spread throughout Brazil and more than 60 countries ^2^ . As P.1 emerged after a period of rapid genetic diversification ^15^, it has accumulated 17 non-synonymous mutations, ten of which are located in the S protein ^13,15^, of which K417T: E484K: N501Y are of biological importance ^16,17^ . Recent studies have reported that P.1 lineage presents an increase in ACE2 receptor affinity due to the presence of the N501Y mutation ^16,18^, which significantly impacts its transmissibility rate, shown to be 1.4-2.2-fold higher than the wild type strain ^13^ . Moreover, P.1 also exhibited a reduction of anti-RBD antibody neutralization ^19^, and it has been implicated in breakthrough infections of vaccinated individuals as well as reinfections ^20,21,22^ . Although several studies documented P.1’s increased transmissibility ^15,23^ and immune evasion ^20,22,24–26^, little is known about its association with the severity of COVID-19 disease and mortality risk, which is crucial to better understand and mitigate the severe impact of the ongoing pandemic.

In addition to P.1’s emergence and dissemination, Brazil has faced a slow vaccination roll-out, which contributed to the prolonging of the pandemic and the emergence of additional SARS-CoV-2 variants. The immunization campaign started in February 2021, in descending age order. As of July 26, 2021, 131,946,091 doses of COVID-19 vaccine have been administered with 44.11% and 17.3% of the population receiving the first and second doses, respectively ^27^ . Brazil has approved the use of four COVID-19 vaccines: ChAdOx1 nCoV-19 (AZD1222; Oxford-AstraZeneca), CoronaVac (Butantan Institute, São Paulo, Brazil and Sinovac Life Sciences, Beijing, China), BNT162b2 mRNA (Pfizer/BioNTech), and Ad26.COV2.S (Janssen /Johnson & Johnson), corresponding to 46.4%, 40.8%, 9.9%, and 2.9% of the administered doses ^27^, respectively. While most vaccines show high efficacy in preventing severe COVID-19 disease and death ^28–33^, the impact of slow vaccination rates on the circulation and spread of VOCs on the epidemiological profiles at the national and local level is still unclear. Here, we report the rapid spread of the P.1 variant following its introduction and dissemination in São José do Rio Preto, Brazil, and show that this event was associated with a change in the epidemiological profile, with increased numbers of severe COVID-19 cases and deaths, especially in the unvaccinated population.

## Methods

### Study area description

São José do Rio Preto (SJdRP) is in the northeast region of the state of São Paulo (SP), Brazil, with a total of 408,258 inhabitants. One of the largest and most important hospital complexes in the municipality is the *Hospital de Base de São José do Rio Preto* (HB). The HB is a reference health center serving more than 2 million inhabitants of the 102 municipalities belonging to the 15th Regional Health Division (RHD XV), headquartered in SJdRP. The HB complex is one of the leading in COVID-19 care and treatment centers in SP state, having the second largest COVID-19 ICU in Brazil, with more than 180 beds and having received more than 5,700 admissions so far. Besides, since the beginning of the COVID-19 pandemic, HB is the main responsible for SARS-CoV-2 diagnosis for the local population. The hospital is linked to the *Faculdade de Medicina de São José do Rio Preto* (FAMERP), an educational facility where *Laboratório de Pesquisas em Virologia* (LPV) is located and where this research was conducted.

### Moving average analysis

A time-trend analysis was performed using a seven-day moving average of notified cases, severe cases, and deaths related to COVID-19 in the RHD XV from March 2020 to May 2021. The data were retrieved from the Public Health System and received from the Reporting Disease Information System (SINAN), using mild respiratory syndrome (e-SUS) and severe acute respiratory syndrome (SRAG) cases databases.

### Samples and molecular investigation

Nasopharyngeal swab samples of residents of SJdRP and nearby cities tested at HB by molecular diagnosis in the COVID-19 routine diagnosis with a positive diagnostic for COVID-19 were obtained between October of 2020 and June of 2021. The total RNA was extracted from 140 µL of nasopharyngeal swab samples, using QIAamp Viral RNA Mini Kit (QIAGEN, Hilden, Germany), following the manufacturer’s instructions. SARS-CoV-2 RNA investigation was performed by one-step real-time polymerase chain reaction (RT-qPCR) using primers and probes targeting the envelope (E), the nucleocapsid (N) region of the SARS-CoV-2 genome, and the human RNAse P according to GeneFinder COVID-19 Plus RealAmp Kit (OSANG Healthcare, KOR) (the manufacturer does not provide sequences of the primers and probe) ^34^ . The RT-qPCR was conducted in a QuantStudio 3 Real-Time PCR System (Thermo Fisher Scientific, USA) with the following conditions: 50°C for 20 minutes for the reverse transcription, 95°C for five minutes for denaturation, followed by 45 cycles of denaturation at 95°C for 15 seconds, and annealing at 58°C for 60 seconds The results were analyzed in QuantStudio 3 software v1.5.1 (Thermo Fisher Scientific, USA) and were interpreted as cycle quantification value (Cq) less or equal 40 as positive and Cq more than 40 as negative. Positive and negative controls, which are included in the GeneFinder COVID-19 Plus RealAmp Kit (non-infectious DNA plasmids coding for the SARS-CoV-2 E gene and N gene), were used in the assay.

The study was approved by the Ethics Committee of Faculdade de Medicina de São José do Rio Preto Institutional Review Board (IRB) (protocol number: CAE# 31588920.0.0000.5415).

### Whole-genome sequencing

Whole-genome sequencing was performed using Next-Generation Sequencing (NGS) technology. The cDNA synthesis, whole-genome amplification, and library preparation were carried out following the instructions provided by Illumina CovidSeq Test (Illumina Inc, USA) and QIAseq SARS-CoV-2 Primer Panel (Qiagen, USA). The quality and size of the libraries were verified by Agilent 4150 TapeStation (Agilent Technologies Inc, USA). Libraries were pooled in equimolar concentrations, and the sequencing was implemented on the Illumina MiSeq System (Illumina Inc, USA), using MiSeq Reagent Kit v2 (read length of 2 x 150 bp) (Illumina Inc, USA).

### Genome assembling and lineage analyses

The quality of FASTQ sequencing data was checked using FastQC software v0.11.9 (http://www.bioinformatics.babraham.ac.uk/projects/fastqc), and trimming was performed in Geneious Prime v. 2021.1 (https://www.geneious.com/), using the plugin BBDuk v. 37.25, to remove primer sequences, adapters, and low-quality bases. A minimum Phred score of Q30 ^35^ and a minimal read length of 75 base pairs (bp) were used. The cleaned paired-end reads were mapped against the hCoV-19/Wuhan/WIV04/2019 (EPI_ISL_402124) genome, available at EpiCoV in GISAID database ^36^ (https://www.gisaid.org/), considering a minimum 50 bp overlap, minimum identity of 90% and maximum mismatches of 10% per read ^37^ . SNPs were identified using default settings and minimum coverage of five reads per site. The genomes were submitted to Pangolin COVID-19 Lineage Assigner Tool ^2^ to confirm the variant classification (https://pangolin.cog-uk.io/).

### Geoprocessing

A database was created with SARS-CoV-2 identified lineages, location/municipality origin, and month of the collected sample. Shapefiles of the areas were downloaded, and maps were created using the QGIS software version 3.8.2 (http://qgis.org).

### Phylogenetic and evolutionary analyses

The datasets used for the phylogenetic analysis included 780 Brazilian SARS-CoV-2 Brazilian complete genomes sequences and the reference genome hCoV-19/Wuhan/WIV04/2019 (EPI_ISL_402124) retrieved from the GISAID database (https://www.gisaid.org/) (Supplementary Table 6). The nucleotide sequences were aligned using MAFFT multiple sequence alignment software version 7.271 ^38^ . Time-scale phylogenetic trees using the Maximum-likelihood (ML) method were reconstructed in IQ-TREE v. 2.0.3 ^39^, using the best-fit model of nucleotide substitution, according to Bayesian Information Criterion (BIC), inferred by ModelFinder application ^40^ . The reliability of branching patterns was tested using Aproximate likelihood-ratio test (aLRT) ^41^ . To investigate the temporal signal from the ML tree, we regressed root-to-tip genetic distances against sample collection dates using TempEst v 1.5.1 (http://tree.bio.ed.ac.uk) ^42^ .

### Epidemiologic analyses

Estimation of the effective reproductive number (R_eff_) for SARS-CoV-2 over the study period was carried out using the package EpiEstim ^43^ in R version 4.0.4 ^44^ . Detailed methods can be found in the documentation of the EpiEstim package. We fit the time-varying Reff assuming a parametric serial interval with mean and standard deviation of five days ^45^ using a 21-day sliding window. To assess potential shifts in severity, we calculate the percentage of all cases that are severe for all age groups and for those 70 and older. We compare the proportion of deaths, non-severe, and severe cases by age to look for qualitative shifts in age distributions due to changing transmissibility or virulence of the changing variants and any potential vaccine effects. To assess the magnitude of increases in deaths as a function of P.1 prevalence, we fit both Poisson and Negative Binomial models to calculate incidence rate ratios for death by age adjusting for the count of tests performed on that day. Finally, we include an index of social mobility ^46^ to track time spent away from the cellphone’s predominant nighttime location.

### Vaccination efficacy

For each age groups, the proportion of cases, severe cases, and deaths in that age group relative to people aged under 60 (the age cutoff for the vaccine rollout) was calculated (a) for the period up until the start of the vaccine rollout for that age group, and (b) for the period starting from two weeks after the end of vaccine rollout for that age group and ending on 29 May 2021. For example, if there were 60% as many deaths in people aged 90+ compared to people aged under 65 before the vaccine rollout began, and 30% as many deaths in people aged 90+ compared to under 65 after the vaccine rollout ended, then the change in proportion of deaths would be –50%. Coverage was calculated as the number of doses divided by target population size divided by two (since two doses are required for full coverage). An ordinary least squares fit was performed using Python 3.8 (https://www.python.org/), stats models v0.12.2 (https://www.statsmodels.org/stable/index.html) to calculate the slope of best fit between the vaccine coverage and the change in proportion of each of cases, severe cases, and deaths, with the intercept fixed at zero.

## Results

### COVID-19 samples characterization

As of July 26, 2021, São José do Rio Preto (SJdRP), a medium-sized city in the Northwest region of São Paulo state, Brazil (Fig. 1a,b,c), has the third-highest number of confirmed COVID-19 cases in the state of São Paulo, reaching 89,332 cases and 2,639 deaths, with a case fatality rate of 3%^46^ . A seven-day moving average of COVID-19 cases showed an increase in hospitalized cases and deaths from February 2021 to April 2021, compared to a year ago (Supplementary Fig. 1). These numbers raised concerns for the epidemiological landscape in SJdRP and the surrounding region. To better understand the events underlying the circulation, transmission, and evolutionary dynamics of SARS-CoV-2, extensive genomic surveillance was implemented starting in October 2020, using nasopharyngeal swabs samples obtained from patients diagnosed with COVID-19 at our reference hospital, Hospital de Base (HB).

**Fig. 1:**
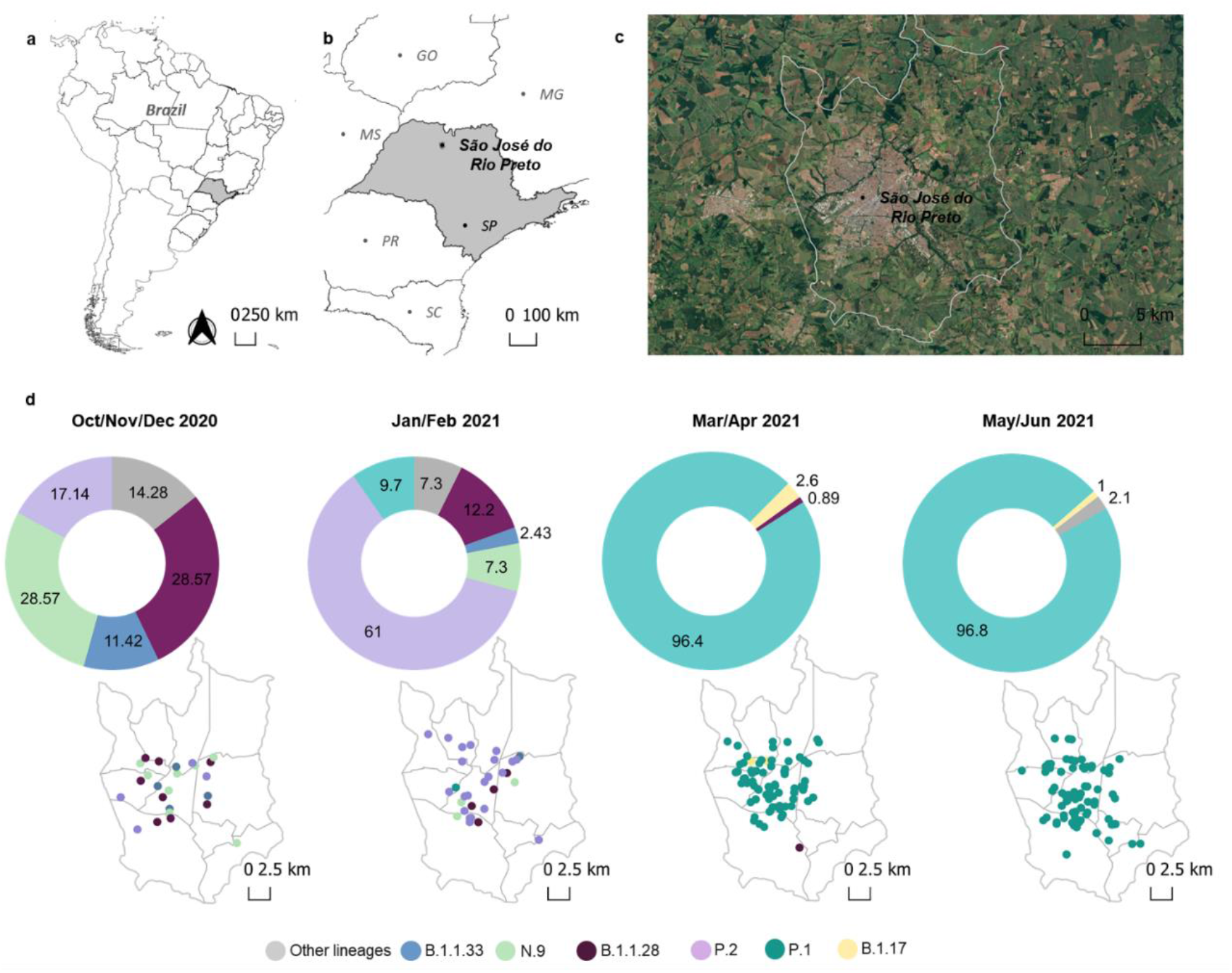
Geographic and temporal distribution of SARS-CoV-2 variants detected in the study area of São José do Rio Preto, São Paulo, Brazil, from October 2020 to June 2021 by genomic surveillance. **a**, Geopolitical map of Brazil and São Paulo state highlighted in grey. **b**, Map of São Paulo state (in grey) indicating the municipality of São José do Rio Preto (SJdRP), located in the Northwest region. **c**, Satellite image of SJdRP. **d**, Prevalence and distribution of SARS-CoV-2 variants detected in SJdRP from October 2020 to June 2021 by genomic surveillance.

From October 2020 to June 2021, a total of 306 SARS-CoV-2 complete genomes were generated at the *Laboratório de Pesquisas em Virologia* (LPV), part of the Corona-Ômica network (http://www.corona-omica.br-mcti.lncc.br), at FAMERP, from samples obtained in SJdRP (88.9% of the sequences) and 15 neighboring municipalities (11.1% of the sequenced genomes) (Supplementary Table 1). Samples were obtained from 165 female (54%) and 141 male patients (46%), with an average age of 43 years (age range of eight months old to 91-years-old).

### Lineage classification of SARS-CoV-2 genomes

SARS-CoV-2 viral lineages (sequences obtained in this study and available at GISAID database) were classified using Pangolin ^2^ showing that the most prevalent lineages circulating in SJdRP were classified into B.1.1.28 (n=16, 5.6%), N.9 (n=13, 4.6%), P.2 (n=31, 11%), P.1 (n=205, 72.4%) and others (which includes B.1, B.1.1, B.1.2, B.1.1.373, B.1.1.33, B.1.1.332, B.1.1.7 and P.1.1(n=18, 6.3%)) (Supplementary Tables 2 and 3). The variant prevalence changed over time, from seven lineages co-circulating in October and November 2020, with a predominance of N.9 (28.5%) and B.1.1.28 (28.5%) to eight lineages in January and February 2021. Additionally, a shift in the prevalence of variants was observed, represented by a fourfold decrease in N.9 frequency (7.3% of sequenced genomes) and an increase of P.2 variant frequency (61% of sequenced genomes). Moreover, in January 2021, P.1 lineage was first detected and contributed to changing the landscape of circulating variants in SJdRP (Fig. 1d) and surrounding cities (Supplementary Fig. 2, Supplementary Tables 4 and 5) for the subsequent months. Critically, following P.1 introduction, a rapid increase in prevalence was observed, reaching more than 96% of the sequenced genomes from March to June, thus replacing P.2 and other lineages (Fig. 2). In April 2021, VOC B.1.1.7 was first detected in SJdRP, and despite its low frequency, it was the only variant identified in May and June together with P.1.1 in addition to P.1 (Fig. 1d). Similarly, a complete replacement of other variants by P.1 was also observed in neighboring municipalities of SJdRP, administered by the Regional Health Division (RHD XV), for which SJdRP serves as the main health center (Supplementary Fig. 2).

**Fig. 2:**
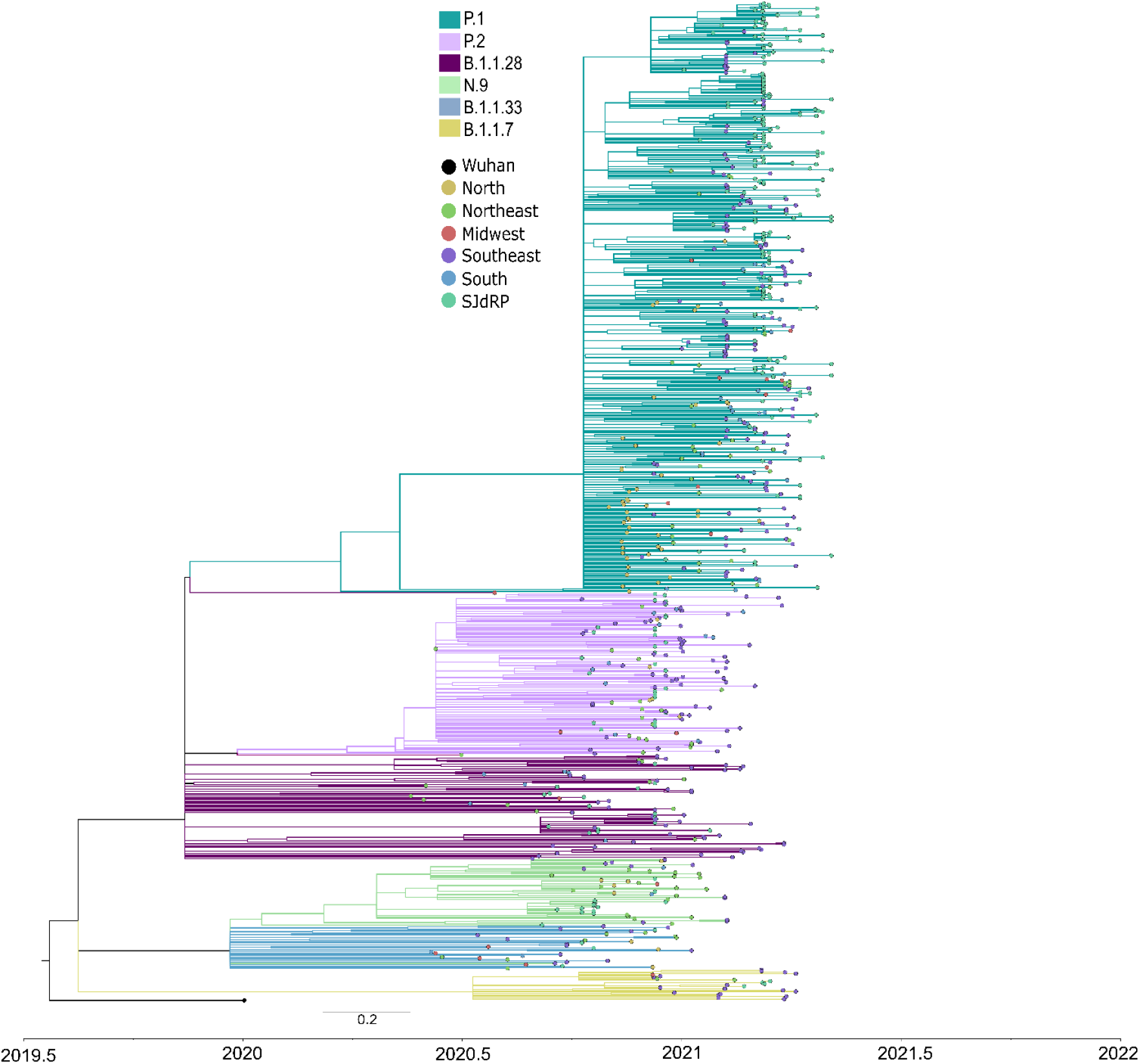
Maximum likelihood tree of SARS-CoV-2 based on complete genome sequences from São José do Rio Preto and all Brazilian regions. Phylogenetic tree reconstructed using GTR+F+R2 as nucleotide substitution model. The reliability of branching patterns was tested using Approximate Likelihood-ratio test (aLRT). The analysis involved 272 complete genome sequences from SJdRP and 508 from five Brazilian regions. The analysis was conducted in IQ-TREE v. 2.0.3, and the final tree was visualized and edited in FigTree v.1.4.4.

### SARS-CoV-2 P.1 variant introduction and clade replacement

We performed a maximum likelihood phylogeographic analysis using a dataset composed of 780 Brazilian complete genome sequences of B.1.1.28, P.1, P.2, N.9, B.1.1.33, and B.1.1.7 variants, being 272 sequences from SJdRP, of which 262 were obtained in this study and 10 were sequences retrieved from the GISAID database (Supplementary Table 6). Our genomic surveillance indicates that multiple introduction events of different SARS-CoV-2 lineages occurred in SJdRP from several Brazilian regions (Fig. 2). The B.1.1.28 variant (n=16), the ancestral lineage of P.1 and P.2, was introduced in SJdRP several times over at least five months. The most prevalent lineages, P.1 (n= 205) and P.2 (n= 31) formed a well-supported group (aLRT= 99.8 and 92.5, respectively), including sequences from four Brazilian geographic regions, further supporting the notion of repeated strain introductions into the area (Fig. 2).

Due to a large number of SARS-CoV-2 P.1 sequences detected in our genomic surveillance, we performed a more targeted phylogenetic analysis of all sequences sampled solely from SJdRP and the surrounding area (Supplementary Fig. 3, Supplementary Tables 7 and 8). P.1 sequences formed a single well-supported group (aLRT= 100) clustered most closely with B.1.1.28 sequences, the parental lineage of P.1 (Fig 3). The first detection of P.1 strains in SJdRP was on 26 January 2021, followed by a rapid increase in its prevalence over other variants. The Fig. 3 shows the dominance of P.1, which displaced all the other variants circulating in SJdRP in less than two months (from March 2021). Root-to-tip analyses confirmed that P.1 was the most divergent variant (Fig. 3). Interestingly, P.1 samples collected at the same period displayed different divergence patterns, which could indicate a diversification process in the P.1 variant circulating in SJdRP (Fig. 3).

**Fig. 3:**
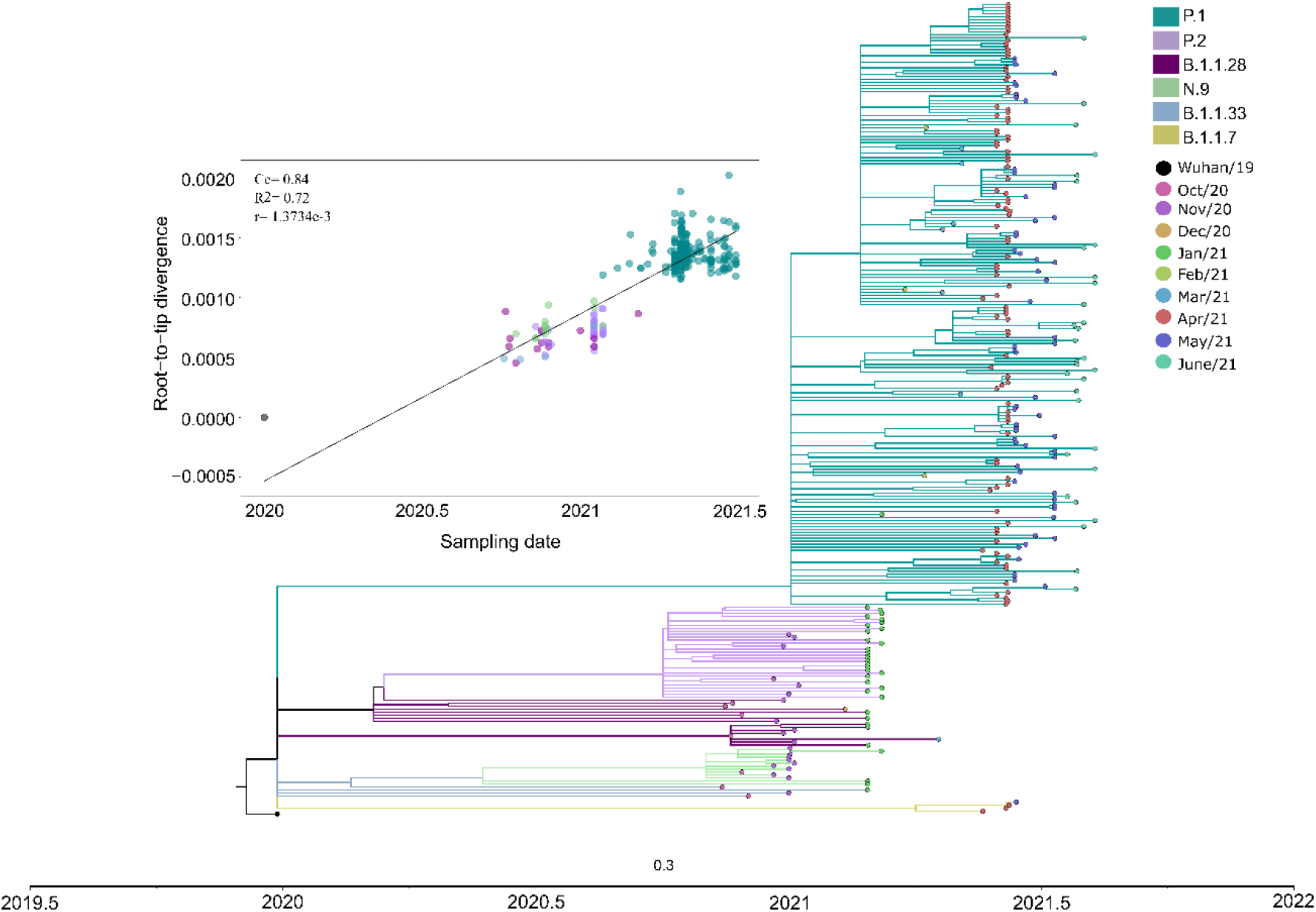
Maximum likelihood tree of SARS-CoV-2 based on complete genome sequences from São José do Rio Preto according to sampling month. The Maximum-Likelihood tree was reconstructed based on the GTR+F+R3 nucleotide substitution model. The reliability of branching patterns was tested using the Approximate Likelihood-ratio test (aLRT). The analysis involved 272 complete genome sequences. The analysis was conducted in IQ-TREE v. 2.0.3, and the final tree was visualized and edited in FigTree v.1.4.4. Correlation between the sampling date of the most prevalent SARS-CoV-2 lineages detected in SJdRP and their genetic distance from the root (hCoV-19/Wuhan/WIV04/2019 - EPI_ISL_402124) based on the Maximum Likelihood phylogenetic tree (Correlation coefficient (Cc)= 0.84; R^2^ = 0.72, substitution rate (r)= 1.3734e-3).

### Epidemiologic profile of the introduction of SARS-CoV-2 P.1 variant

Deaths showed a marked increase as variant P.1 became dominant (Fig. 4a), increasing by 162% (95% CI: 127, 214) when comparing July-September 2020 to March-April 2021.While the over 70 years of age group accounted for most deaths across the epidemic, shifts were seen from older to younger age groups as P.2 waned and P.1 dominated (Fig. 4b).

**Fig. 4:**
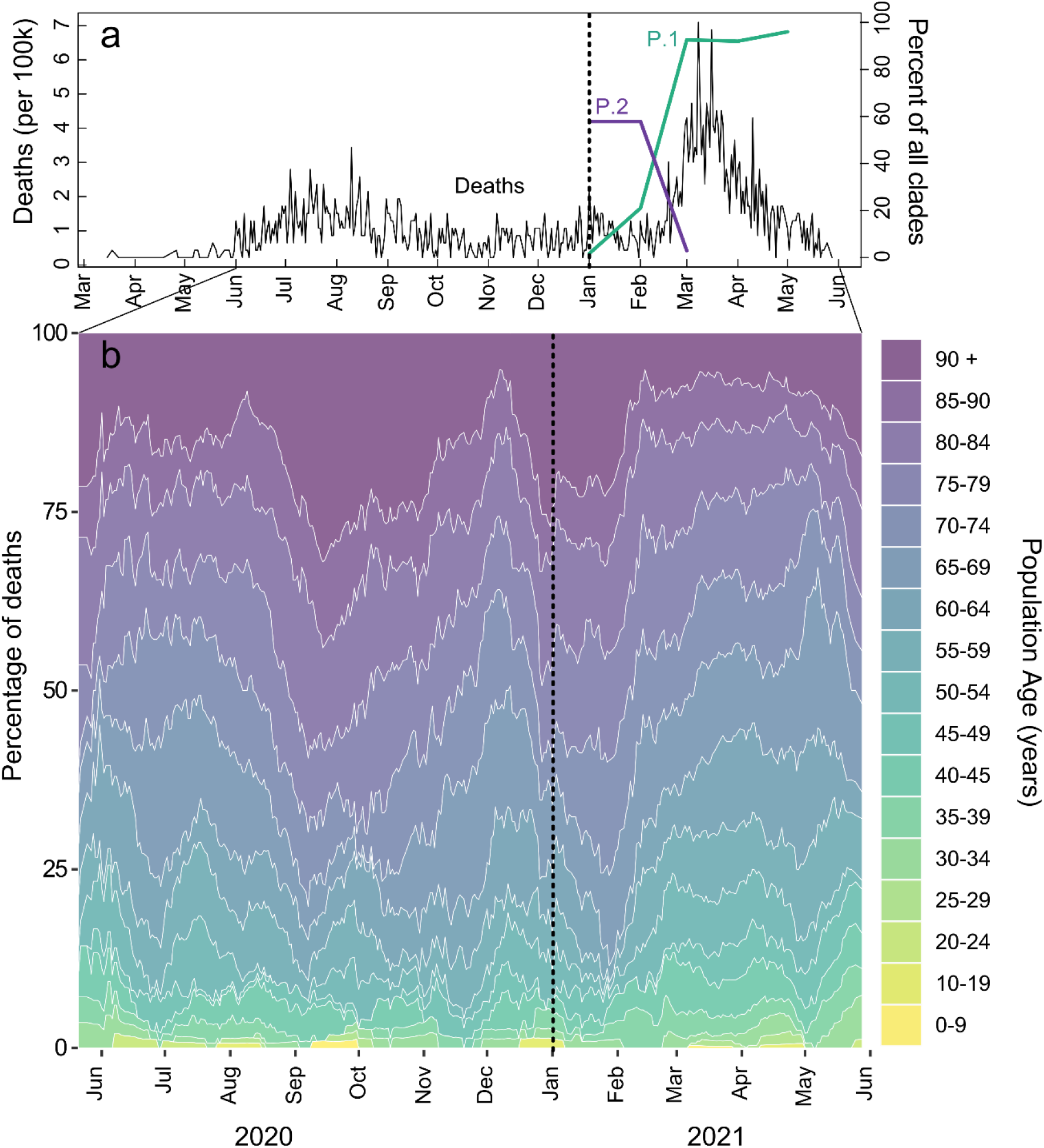
Increase of deaths associated with SARS-CoV-2 P.1 lineage introduction and spread in São José do Rio Preto. **a**, COVID-19 deaths per 100,000 inhabitants from March 2020 to June 2021 and proportion of P.1 and P.2 clades. **b**, Percentage of COVID-19 deaths by population age.

Cases per 100,000 fluctuated across March 2020 to June 2021 with corresponding estimates of R_eff_ (Fig. 5a, b). Peaks in cases and R_eff_ were seen in July-August 2020, November 2020, January 2021, and the highest peaks in March and April 2021, followed by reduced social isolation index (Fig. 5a, b) and the rise in dominance of P.1 from March 2021. Non-severe cases were nearly 10-fold higher than severe cases during most of the time of the pandemic, however, an increase of 127% (95% CI: 105, 152) of severe cases was seen in March 2021, corresponding to the rise of P.1 (Fig. 5c, d). Proportionally, the largest increase in severe cases was seen in those under 70 years old (109% increase comparing January to March 2021; 95% CI: 78,149) as opposed to those over 70, which had a 19% (95% CI: 7, 33) increase (Supplementary Fig. 4). Negative binomial regressions were far superior by AIC than Poisson models. Incidence rate ratios (IRRs) for death ranged from 1.3 (95% CI: 0.63, 2.62) for those aged 30-34 to 2.36 (95% CI: 1.68, 3.49) for those aged 45 to 49 (Fig. 6a). Correspondingly, IRRs were higher overall for those under 70 versus over 70 (1.75, 95% CI: 1.34, 2.38 versus 1.54, 95% CI: 1.42, 1.68), indicating that people aged from 45 to 69 present higher mortality rate due to P.1 infection, likely due to less vaccination coverage by the time of the study (Fig. 6a). Additionally, daily surveillance of the available Intensive Care Unit (I.C.U.) beds for COVID-19 patients in SJdRP revealed that the occupancy rate reached 100% only in a few days in March 2021, corresponding to the increase of P.1 prevalence and the number of severe cases (Supplementary Fig. 5). Nevertheless, as no collapse in the healthcare system was verified, most deaths observed from March 2021 are probably linked to a more severe outcome of P.1 infection than lack of proper medical care.

**Fig. 5:**
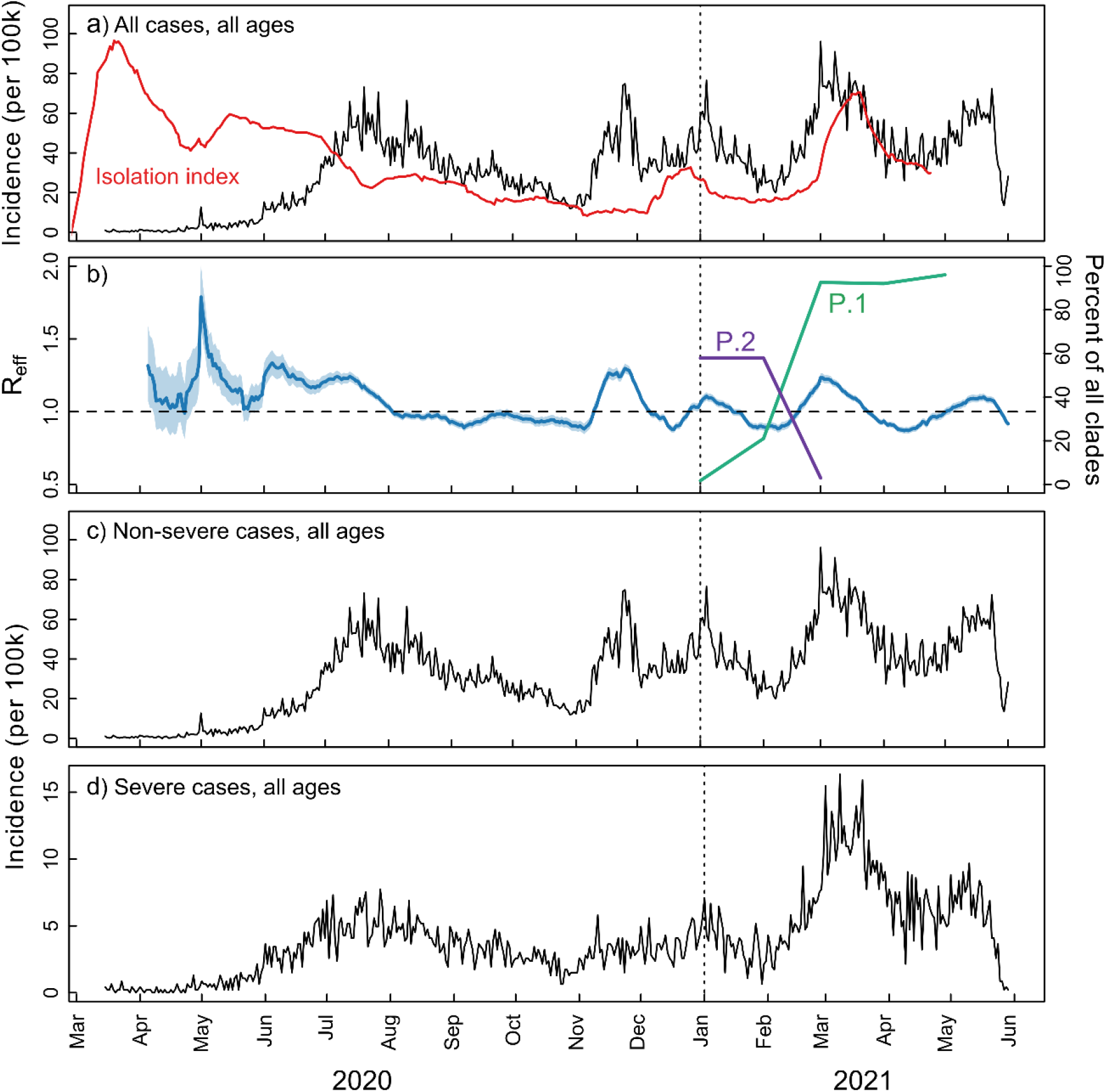
COVID-19 incidence per 100,000 inhabitants. **a, c, d**, COVID-19 incidence per 100,000 inhabitants for total, non-severe, and severe cases, respectively. **b**, Estimates of R_eff_ overtime for all COVID-19 cases as well as the percentage of P.1 and P.2 of all clades.

**Fig. 6:**
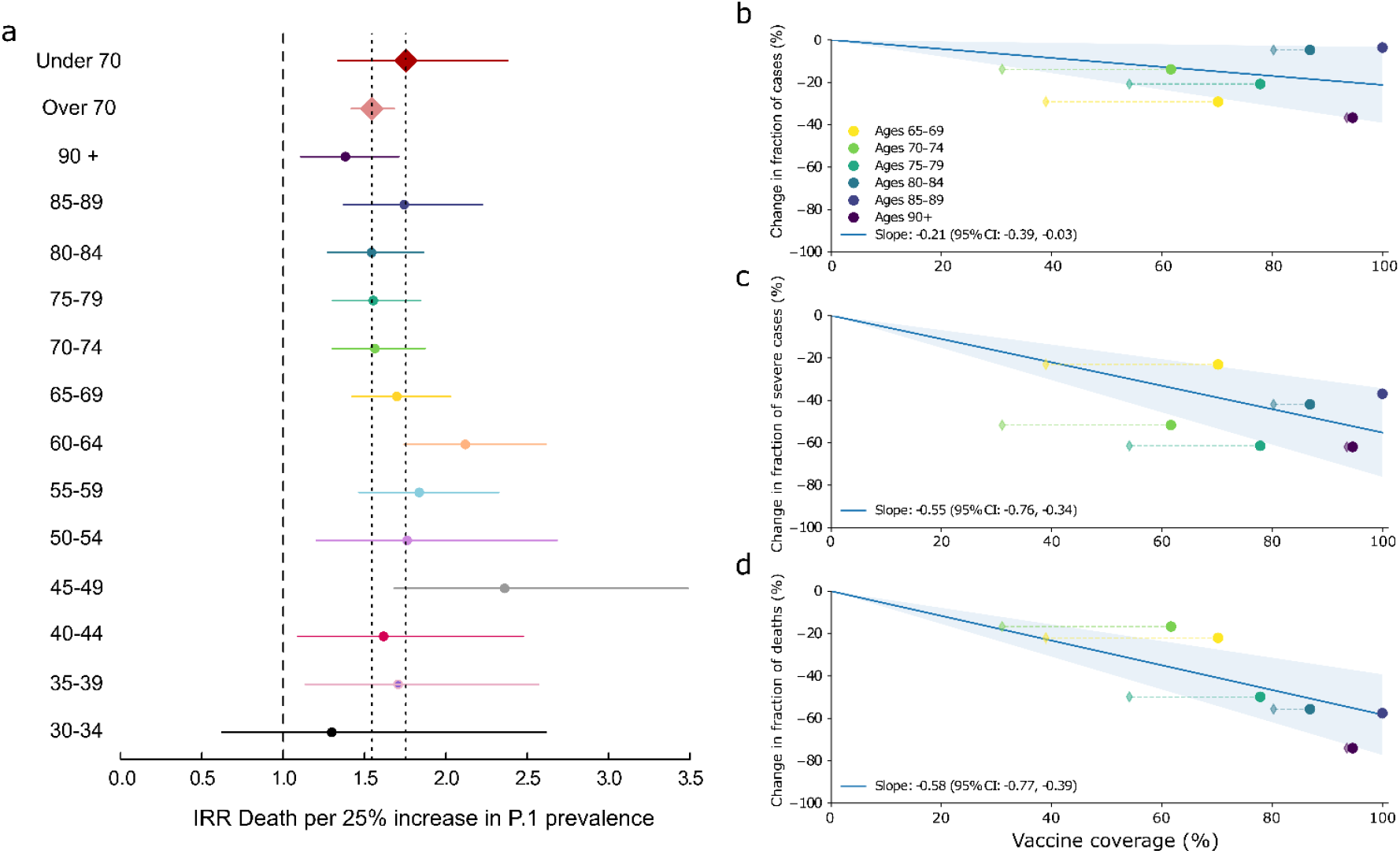
Increased mortality risk associated with P.1 introduction and estimated impact of vaccination on transmission, severity, and mortality. **a**, Incidence rate ratio (IRR) for death for each 25% increase in P.1 prevalence, adjusted for per-day numbers of tests performed. **b**, Change in the proportion of cases in each age group from before versus after vaccination. Circles show effective vaccine coverage (total doses divided by twice the population), while diamonds show the percentage of people who have received both doses. **c, d**, Change in proportions of severe cases and deaths, respectively.

### Effects of COVID-19 vaccination

CoronaVac and AstraZeneca vaccines were rolled out to people in descending age order, beginning with those aged over 90 years-old on 2 February 2021. To estimate the impact of vaccination, we compared the proportion of cases, severe cases, and deaths in each age group before the rollout (for that age group) versus two weeks after the rollout onwards. A line with a fixed intercept was fit to these changes in proportion, such that a slope of –1 corresponds to one case, severe case, or death being averted per person vaccinated. As shown in Fig. 6b, vaccination was associated with a moderate reduction in the number of cases (best-fit slope – 0.21, 95% CI: –0.03, –0.39). However, it was associated with a pronounced reduction in severe cases (–0.55, 95% CI: –0.34, –0.76) and deaths (–0.58, 95% CI: –0.39, –0.77) (Fig. 6c, d). A slope of less than –1 for deaths, for example, would imply that more than one death is averted per person vaccinated; this would be the case if, for example, people who are vaccinated not only have significantly lower mortality rates if infected but if they also have lower rates of transmission, which preferentially reduces infection rates among unvaccinated people in the same age group (herd immunity).

## Discussion

For more than eighteen months, the world has lived amid a pandemic, with an alarming number of cases and deaths and devastating consequences in all sectors of society. Nevertheless, the current situation in some countries is encouraging, with the number of cases and deaths decreasing rapidly following mass vaccination. Unfortunately, Brazil remains to be an epicenter of SARS-CoV-2 transmission due to the persistent absence of control measures and low vaccination rates^47^ . From the first recorded case in February 2020 to July 2021, Brazil recorded more than 19.5 million laboratory-confirmed cases with more than 549 thousand deaths ^47^ . Here, using a combination of whole genome sequencing and epidemiological analysis, we show that the introduction, rapid spread, and dominance of SARS-CoV-2 P.1 lineage in SJdRP was associated with an increase of severe COVID-19 cases and deaths. Moreover, the mortality risk related to P.1 infection largely depends on several factors, including age, underlying conditions, and lack of vaccination.

The P.1 lineage emerged in early November 2020 in Manaus, the capital of Amazonas state, and it quickly spread to other Brazilian states, mainly to the South-eastern region ^13^ . In early February 2021, a total of 41 complete-genomes of SARS-CoV-2 P.1 lineage were available in the GISAID database (4.8% from Pará, 12.1% from São Paulo, and 82.9% from Amazonas states). More than 49 thousand P.1 genomes have been deposited worldwide, of which 27.14% were contributed from Brazil ^48^ . Our genomic surveillance documented the first detection of P.1 lineage in SJdRP on January 26, 2021, only two months after its detection in Manaus. Following its introduction in the region, this variant quickly became the most dominant lineage, outplacing eight circulating lineages of SARS-CoV-2. These observations mirror trends reported in Manaus and other Brazilian regions ^13,49,50^, attributing its rapid spread and dominance to its higher transmissibility rate and greater viral fitness ^13^ . This is concerning as clade replacement can maintain disease in populations through successive introductions of new lineages ^51^, especially in countries considered epicenters of the pandemic, as have been shown previously with dengue and Influenza viruses ^52–57^ .

We demonstrate that a clade replacement event driven by the introduction of P.1 lineage was able to change the epidemiological landscape of COVID-19 in the municipality. The detection and prevalence of P.1 led to a prominent peak in the total cases and deaths recorded in SJdRP. Interestingly, there was a 109% increase in the number of COVID-19 severe cases of individuals under 70 years, while in the group of individuals over 70 years, there was only a 19% increase of severe COVID-19 cases. This finding is most likely due to the impact of recent vaccinations of older individuals, resulting in significant reductions in both severe disease and mortality. Our findings of the incidence rate ratio of deaths by COVID-19 infection after P.1 introduction revealed a higher mortality risk in people under 70 years, which was more accentuated in individuals aged 45 to 64 years old. These results are like other recent work on the P.1 variant, which reported significant increases in the case fatality rate in those under 50 years of age after the introduction of P.1 in the Brazilian state of Paraná immediately to the south of SJdRP ^50^ .

Furthermore, our results highlight the urgency of rapid and thorough vaccination in all age groups. We showed a pronounced reduction in severe cases and deaths in age groups that have received both doses of SARS-CoV-2 vaccines, meaning that for each vaccinated person, more than one death can be averted, not only due to lower mortality risk, but also due to lower transmission rates for unvaccinated people (herd immunity). In fact, studies have been reported that besides the protective effect of vaccines against SARS-CoV-2 variants, mainly in severe cases ^28,29,31,32,58,59^, vaccination can reduce the viral load in infections occurring 12-37 days after the first dose ^60^, therefore suppressing virus spread. Thus, if efficient vaccination measures are taken rapidly, it is possible to lower the transmission rate, and the mortality risk, for all age groups. Moreover, a rapid and efficient vaccination scenario is essential to reduce the virus divergence, and thus, the emergence of new and more concerning SARS-CoV-2 variants can drive new clade replacement events.

## Data Availability

Data availability
All SARS-CoV-2 genomes generated and analyzed in this study are available at EpiCoV database in GISAID (https://www.gisaid.org), and their respective access number are available at Supplementary Material.

## Data availability

All SARS-CoV-2 genomes generated and analyzed in this study are available at EpiCoV database in GISAID (https://www.gisaid.org), and their respective access number are available at Supplementary Material.

## Acknowledgments

We acknowledge Rede Corona-Ômica BR MCTI/FINEP affiliated to RedeVírus/MCTI (FINEP= 01.20.0029.000462/20, CNPq= 404096/2020-4, CNPq fellowship= 382032/2020-9 to C.A.B), FINEP (Grant #01180149-00), Multiuser Laboratory (LMU) at São José do Rio Preto School of Medicine (FAMERP), Brazil, for their support with the use of equipment’s (MiSeq™, Illumina Inc, USA). Funding support is acknowledged from FAPESP-COVID Program (Grant #2020/04836-0 to MLN), JBS Support for COVID-19 Response Research, FAPESP fellowships (2020/07419-0 to GRFC), Fundação Butantan (FAPESP Grant #2020/10127), Capes (Grant #0001), and partly by the Centers for Research in Emerging Infectious Diseases (CREID), “The Coordinating Research on Emerging Arboviral Threats Encompassing the Neotropics (CREATE-NEO)” grant 1U01AI151807 awarded to NV by the National Institutes of Health (NIH/USA). MLN, PR, JPAJ are CNPq Research Fellows. The funders had no role in the design of the study, collection, analyses, or interpretation of data, writing of the manuscript, or in the decision to publish the results.

## Author contributions

C.A.B, L.S, B.M.A., N.V. and M.L.N conceived and designed the study. G.R.F.C., B.C.M., M.M.M., T.M.I.L.S., B.H.G.A.M., L.C.R., F.S.D., A.L.S., V.B.C., C.P., R.A.S., E.E.O.L., V.M.H., N.Z., C.C.P, C. F. E. and M.C.P.P. performed SARS-CoV-2 diagnostic. C.A.B., L.S., G.R.F.C., M.M.M., F.S.P., L.S.U., B.C.M., G.C.D. carried out the genome sequencing. C.A.B. performed genome assembling. A.F.N., A.C.B., M.D.B. and A.F.V., provided the epidemiological data. J.A.C., C.C.K., B.M.A. performed statistical and epidemiological analyses. C.A.B., L.S., J.A.C., C.C.K., B.M.A. carried out data analyses and interpretation. C.A.B., L.S., C.C.K., B.M.A., N.V., M.L.N. wrote the first draft of the manuscript. C.A.B., L.S., C.C.K., B.M.A., N.V., M.L.N. edited and revised the manuscript. H.L.F., P.R, J.P.A.J, B.M.A. N.V. and M.L.N. provided the resources for the survey. All authors approved the final version of the manuscript. All authors had full access to all the data used in this study and had final responsibility for the decision to submit for publication.

