## Supplemental Fig 1-5 for "Effects of SARS-CoV-2 P.1 introduction and the impact of COVID-19 vaccination on the epidemiological landscape of São José Do Rio Preto, Brazil"

**Supplemental files**

**Supplementary Fig 1: Moving average of daily cases from March 2020 to May 2021.**

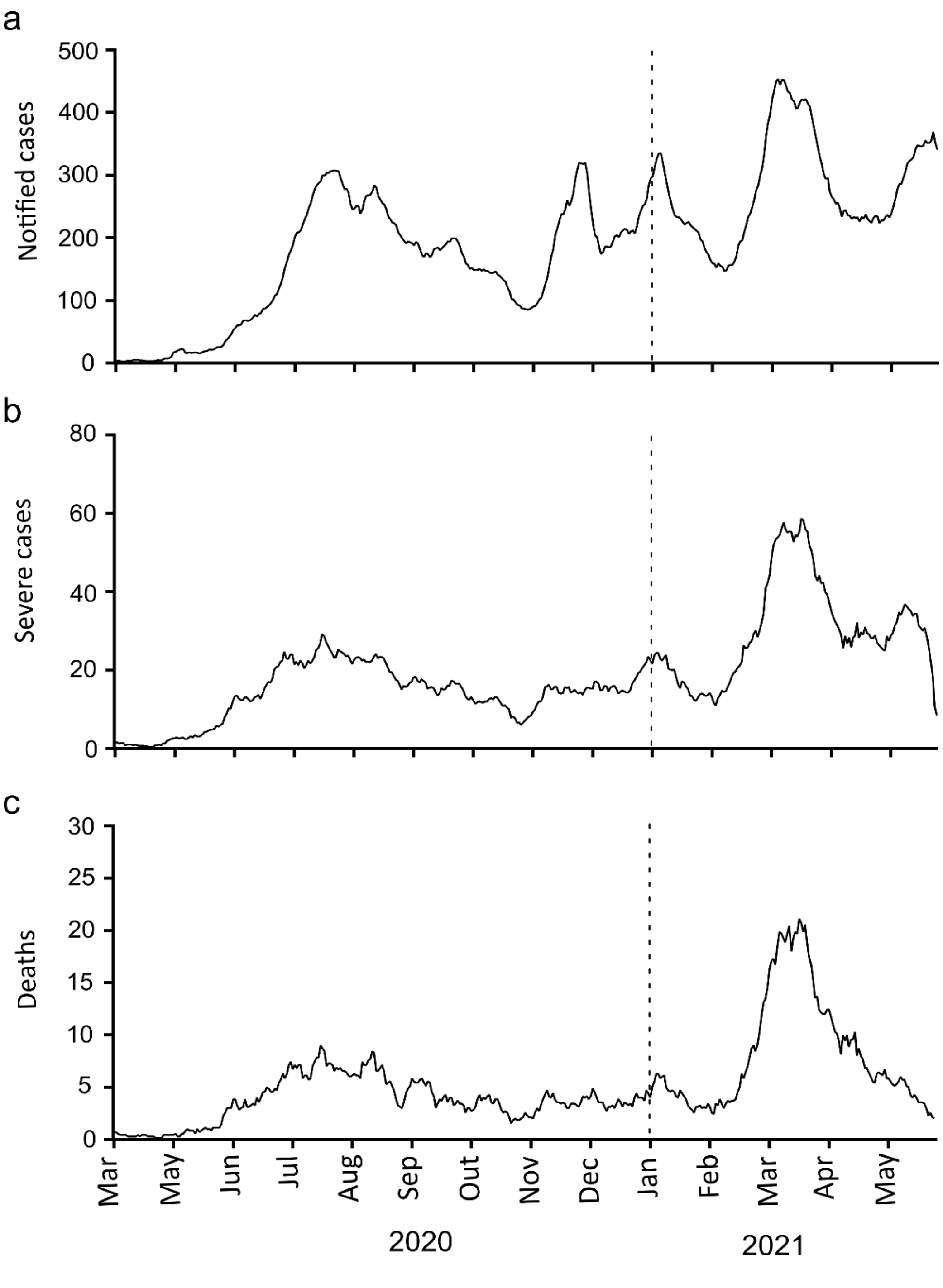


**a,** Moving average of total notified cases. **b,** Moving average of severe cases. **c,** Moving average of deaths in the Regional Health Division XV (RHD XV) of São José do Rio Preto.

**Supplementary Fig 2: Geographic and temporal distribution of SARS-CoV-2 variants detected in the Regional Health Division (RHD XV) of São José do Rio Preto from October 2020 to June 2021 by genomic surveillance.**

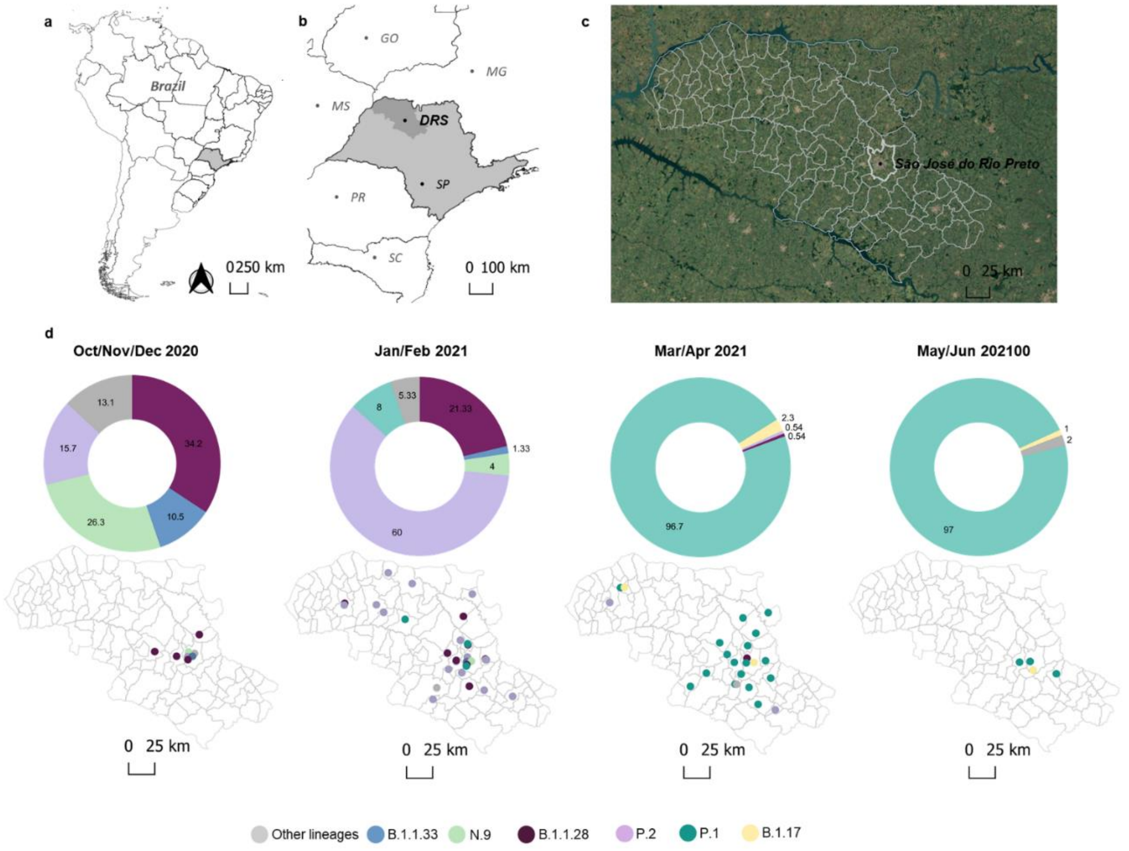


**a,** Geopolitical map of Brazil and São Paulo state highlighted in grey. **b,** Map of São Paulo state (in grey) indicating the Regional Health Division (RHD XV) of São José do Rio Preto (SJdRP), located in the Northwest region. **c,** Satellite image of RHD XV. **d,** Prevalence, and distribution of SARS-CoV-2 variants detected the RHD XV of SJdRP from October 2020 to June 2021 by genomic surveillance.

**Supplementary Fig 3: Maximum likelihood tree of SARS-CoV-2 based on complete genome sequences from the Regional Health Division of São José do Rio Preto.**

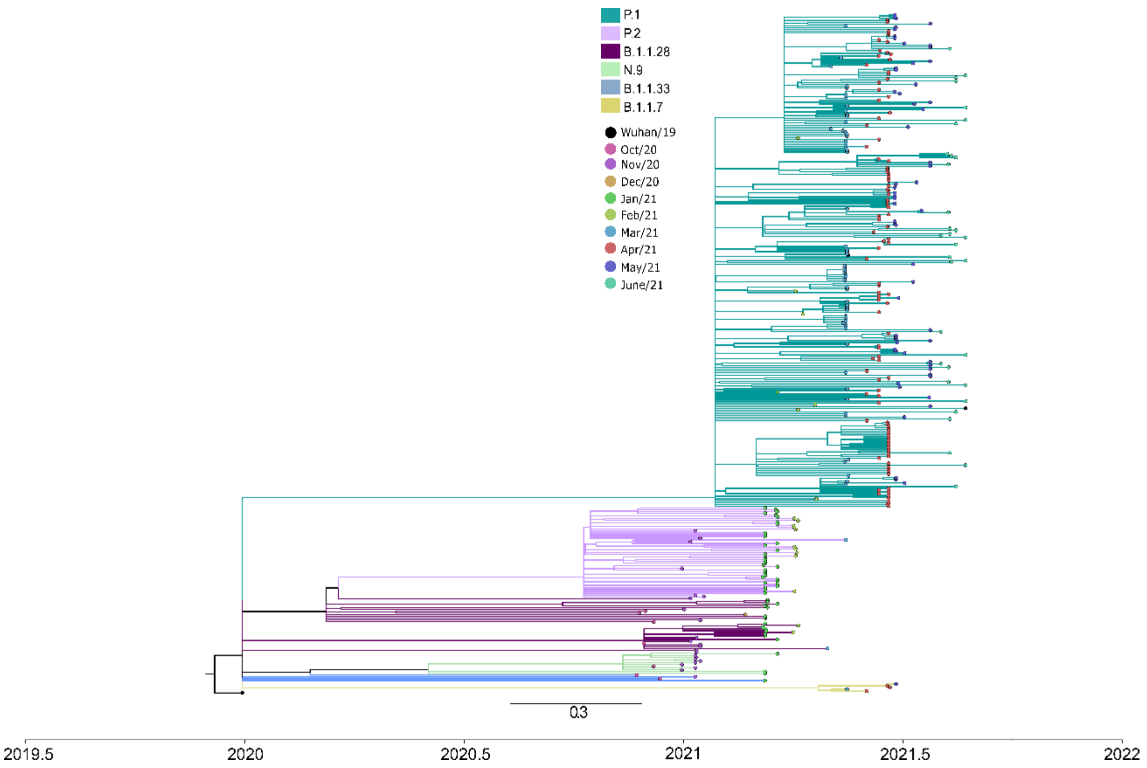


The phylogenetic tree was reconstructed using the Maximum Likelihood method based on the GTR+F+R4 nucleotide substitution model. The reliability of branching patterns was tested using Approximate Likelihood-ratio test (aLRT). The analysis involved 382 complete genome sequences. The analysis was conducted in IQ-TREE (v. 2.0.3), and the final tree was visualized and edited in FigTree v.1.4.4.

**Supplementary Fig 4: Increase of case severity associated with SARS-CoV-2 P.1 lineage introduction and spread in São José do Rio Preto.**

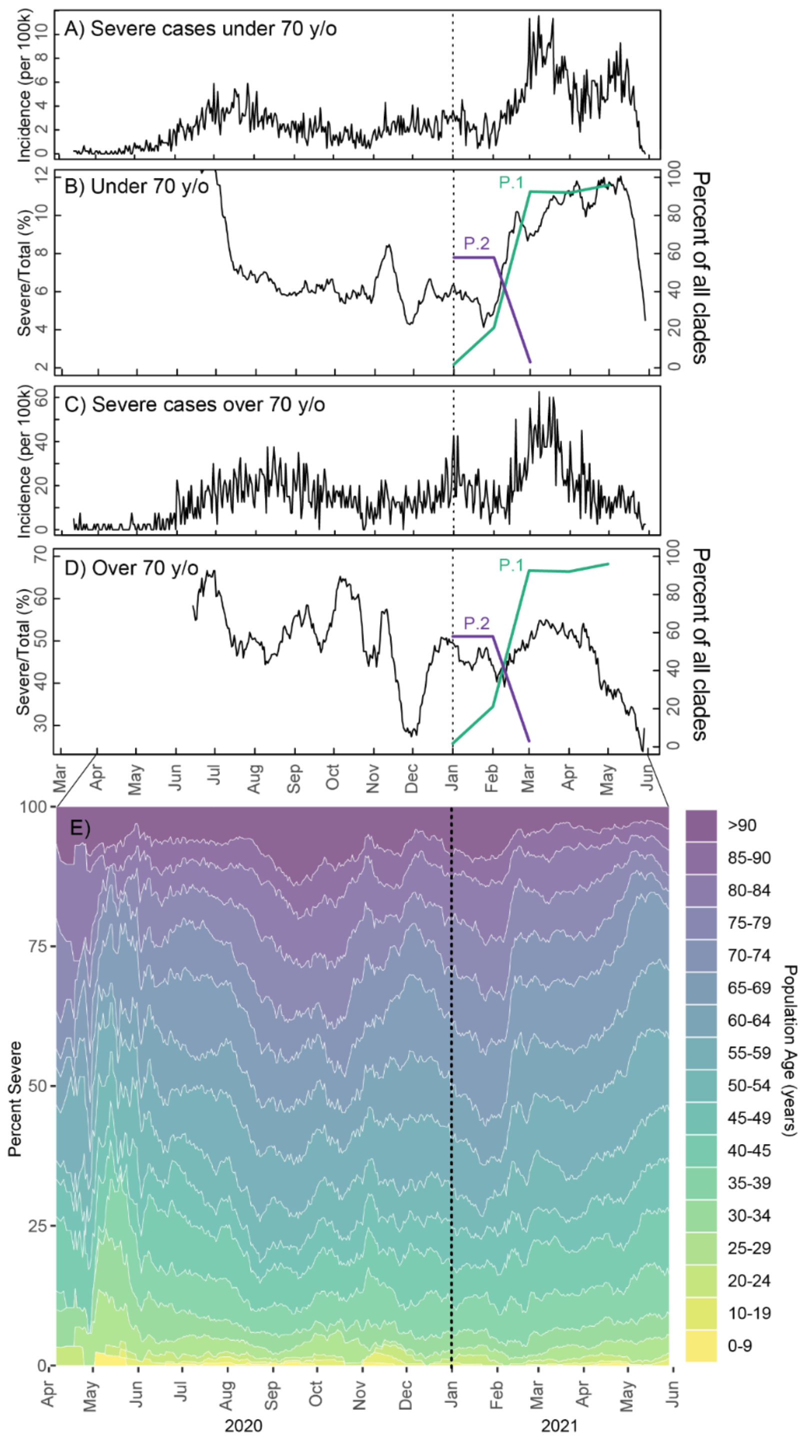


**a, b, c, d,** incidence of SARS-CoV-2 and the proportion of COVID-19 cases that are severe for those under 70 and over 70 years, respectively. **e,** age breakdown of severe cases by age and time.

**Supplementary Fig 5: Percentage of Intensive Care Unit (I.C.U.) beds occupied per month.**

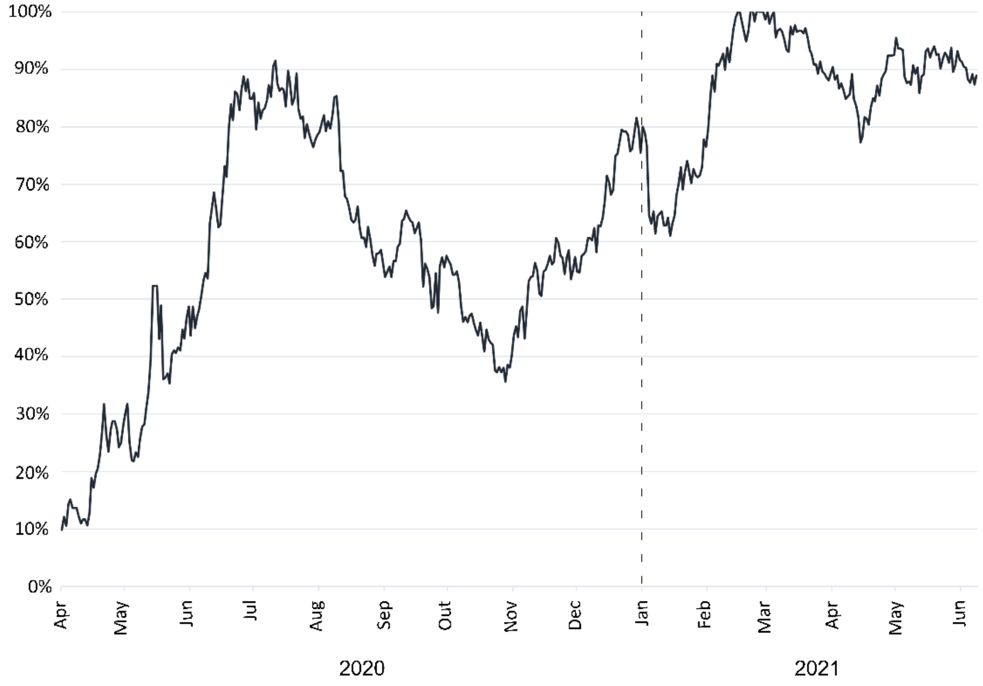


Rate of Intensive Care Unit (I.C.U.) beds occupied by patients diagnosed with COVID-19 in São José do Rio Preto.
